# The transmission dynamics of Norovirus in England: a genotype-specific modelling study

**DOI:** 10.1101/2025.06.13.25329567

**Authors:** Juan F. Vesga, Amy Douglas, Cristina Celma, Edward S. Knock, Marc Baguelin, W. John Edmunds

**Affiliations:** Modelling & Economics Unit, UK Health Security Agency, London, UK; MRC Centre for Global Infectious Disease Analysis and NIHR Health Protection Research Unit in Modelling and Health Economics, Imperial College School of Public Health, London, UK; Gastrointestinal Infections, Food Safety and One Health Division, UK Health Security Agency, London, UK; Enteric Virus Unit, Public Health Reference Microbiology Division, UK Health Security Agency, London, UK; Centre for Mathematical Modelling of Infectious Diseases, London School of Hygiene & Tropical Medicine, London, UK

**Keywords:** Norovirus, Mathematical Modelling, Acute gastroenteritis, Genogroup, Genotype

## Abstract

**Background:** Norovirus is the leading cause of acute gastroenteritis cases in England and worldwide, with diverse co-circulating genotypes. Recent surveillance in England shows GII.17 surpassing GII.4 in prevalence among reported cases, and vaccine candidates targeting multiple genotypes are advancing. However, most transmission models still focus on single-strain dynamics, limiting their ability to assess the role of co-circulating strains on population burden.

**Methods:** We developed an age structured multistrain transmission model that integrates norovirus genotype diversity, waning immunity, and cross-protection within genogroups. We calibrate to case and genotyping surveillance time-series from UKHSA with community-wide age structured incidence estimates and cross-sectional seroprevalence among English children to capture the transmission dynamics of the main co-circulating Norovirus strains in England. Using a calibrated model, we examine the case of an emerging GII.4 variant under different scenarios of transmissibility.

**Results:** We found that on average the current GII.4 strain has an R0 of ∼3.6. We estimate the average number of lifetime Norovirus-caused AGE cases per person to be between 3 and 4, with 60% of children in England experiencing at least one symptomatic episode by the age of four. Our inference analysis suggests that cross-protection within genogroups (between strains of the same genogroup, like GII.4 with other GII), is very limited at conferring protection. Importantly, our modelling suggests that a potential emerging variant would only cause a larger epidemic season if such variant was more transmissible, while equally transmissible new variants would create a re-distribution of co-circulating strains but maintaining the same epidemic size overall.

**Conclusions:** This approach addresses key limitations of single-strain frameworks and offers a more comprehensive understanding of norovirus dynamics, improving the capacity to assess the potential population-level effects of upcoming multivalent vaccine strategies.

## 1. INTRODUCTION

Norovirus is a leading cause of acute gastroenteritis (AGE) globally, accounting for an estimated 3.7 million infections annually in the United Kingdom (UK) alone (Harris et al., 2017). During the 2024/2025 season, 9,289 laboratory-confirmed cases were reported in England up to February 2025 (“National norovirus and rotavirus report, week 9 report: data to week 7 (data up to 16 February 2025) - GOV.UK,” n.d.).Serological studies indicate that over 60% of UK children are exposed to at least one norovirus genotype by the age of four, highlighting significant transmission within paediatric populations (Lindesmith et al., 2023). The virus’s highly contagious nature enables rapid dissemination in enclosed settings such as households, childcare facilities, hospital wards and care homes.

Norovirus is classified into ten genogroups (GI–GX), of which, only GI, GII, and GIV are pathogenic to humans (Chhabra et al., 2019). Within these genogroups, genotypes and variants exhibit extensive genetic diversity, clearly reflected in the recurrent surge of pathogenic variants of GII.4 strain (i.e., genogroup II genotype 4) making it the most prevalent genotype recorded in humans (Farahmand et al., 2022). Phylogenetic studies show that new GII.4 variants emerged approximately every 3–4 years between 1996, namely 1996 Grimsby, 2002 Farmington Hills, 2004 Hunter, 2006 Den Haag, 2009 New Orleans, and 2012 Sydney(Kendra et al., 2021; Parra, 2019), each one replacing the preceding dominant strain and causing widespread outbreaks (Chhabra et al., 2024a). Sydney (SY) 2012 remains the dominant GII.4 variant and the mechanism behind this long persistence is not totally understood, but the existing evidence points to immune imprinting as a plausible explanation(Lindesmith et al., 2022).

Despite GII.4 dominance, the landscape of Norovirus transmission involves co-circulation of strains. In England, GII.17 has replaced GII.4 as the most frequent GII genotype found in molecular surveillance in the season 2024/2025 (53.2% and 28.5%(“National norovirus and rotavirus report, week 9 report: data to week 7 (data up to 16 February 2025) - GOV.UK,” n.d.). This trend has also been observed in Europe and the United States(Chhabra et al., 2024b). Other co-circulating strains have displayed their capacity to produce outbreaks, like GI.3, commonly associated with outbreaks among children(Chiu et al., 2020; Gomes et al., 2024). Although comparatively much less prevalent than GII.4, GI.3 is the most common GI strain in England(“National norovirus and rotavirus report, week 9 report: data to week 7 (data up to 16 February 2025) - GOV.UK,” n.d.)[10].

Vaccine development efforts are advancing, with a number of candidates currently in the pipeline. Among these, two have reached phase III clinical trials, though one trial was suspended due to low efficacy (“HilleVax’s norovirus phase 2b vaccine trial for infants falls short,” n.d.; “UK’s first norovirus mRNA vaccine trial launched | NIHR,” n.d.). The leading vaccine formulations include genotypes GII.4 and GI.1 (Norwalk prototype strain), with some candidates incorporating additional genotypes such as GI.3, GII.17 and GII.3 (Armah et al., 2023).

Transmission models of Norovirus have been developed to explore the basic mechanisms of Norovirus infection and immunity (Lopman et al., 2014; Milbrath et al., 2013; Simmons et al., 2013), analyse localised outbreaks (McMenemy et al., 2023; Vanderpas et al., 2009; Yang et al., 2021), assess transmission patterns post-COVID-19 (Lappe et al., 2023; O’Reilly et al., 2021), and evaluate the potential impact of vaccines (Gaythorpe et al., 2018a; Steele et al., 2016). However, most models assume a single-strain transmission framework, which may limit their applicability given the co-circulation of different genotypes and the multivalent nature of current vaccine candidates.

Here, we propose a novel multistrain transmission model that accounts for the genetic diversity and differential dominance of norovirus genotypes. This approach aims to provide a more comprehensive understanding of the virus’s transmission dynamics and the interaction with host population immunity. The added complexity of our model also allows us to fit it to strain-specific data using a Bayesian evidence synthesis framework(Baguelin et al., 2013). By incorporating genotype-specific parameters, our model seeks to address the limitations of single-strain frameworks while elucidating the immunological mechanisms underlying waning immunity and cross-protection within genogroups.

## 2. METHODS

### 2.1. Definitions

For simplicity, we use a broader definition of “strain” to streamline the tracking of categories in our model. When referring to “genogroup,” we adhere to the strict Norovirus classification. Specifically, Genogroup I (GI) includes genotypes such as GI.3 and other GI types, while Genogroup II (GII) encompasses GII.4 and other GII genotypes. In this context, “strain” refers to either a specific genotype (e.g., GII.4) or a grouped category of genotypes (e.g., other GI types).

Other important clarity is about what we refer in this context as cross-protection and strain-specific immunity. Cross-protection refers to the protective immunity against a genotype conferred by exposure to a genotype of the same genogroup (e.g., immune protection against GII.4, arising from exposure to other-GII strains). In our model, we introduce independent parameters for cross-protection within genogroups (i.e., GI and GII), and assume the absence of cross-protection between genogroups, as has been found in previous studies(A. Ford-Siltz et al., 2021). Strain-specific immunity refers to immunity against the same genotype (“strain”). This immunity in our model is fully protective and its duration is controlled by a waning immunity parameter. This can also vary according to the number of AGE episodes, as explained in *Model Selection* section and **Figure 1**.

**Figure 1.**
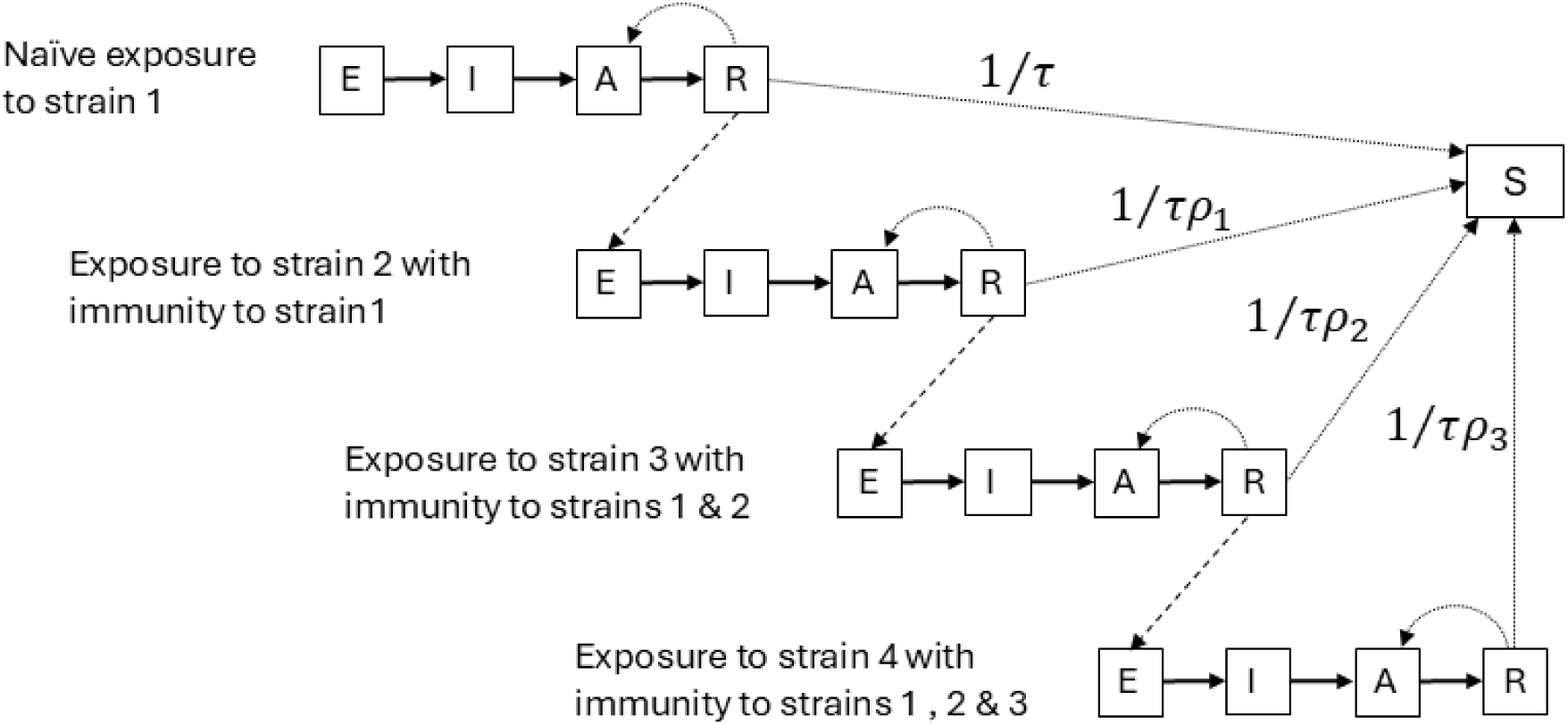
Waning immunity schematic. Sequential exposure to the four simulated strains, with each infection event leading to the subsequent strain infection or waning immunity to full susceptibility. Here τ represent the average waning immunity time (years) and ρ (1 to 3) is a factor increasing the waning time as new strain exposures are accrued. In the model all possible sequences are simulated (e.g., initial GI.3, followed by other GII, etc..).

### 2.2. Mathematical model

We designed an age-structured compartmental, stochastic transmission model of four genotypes of Norovirus (i.e., GI.3, GII.4, other GI genotypes and other GII genotypes). **Figures 1 and 2** describe the details of the multi-strain structure and natural history of disease in the model.

**Figure 2.**
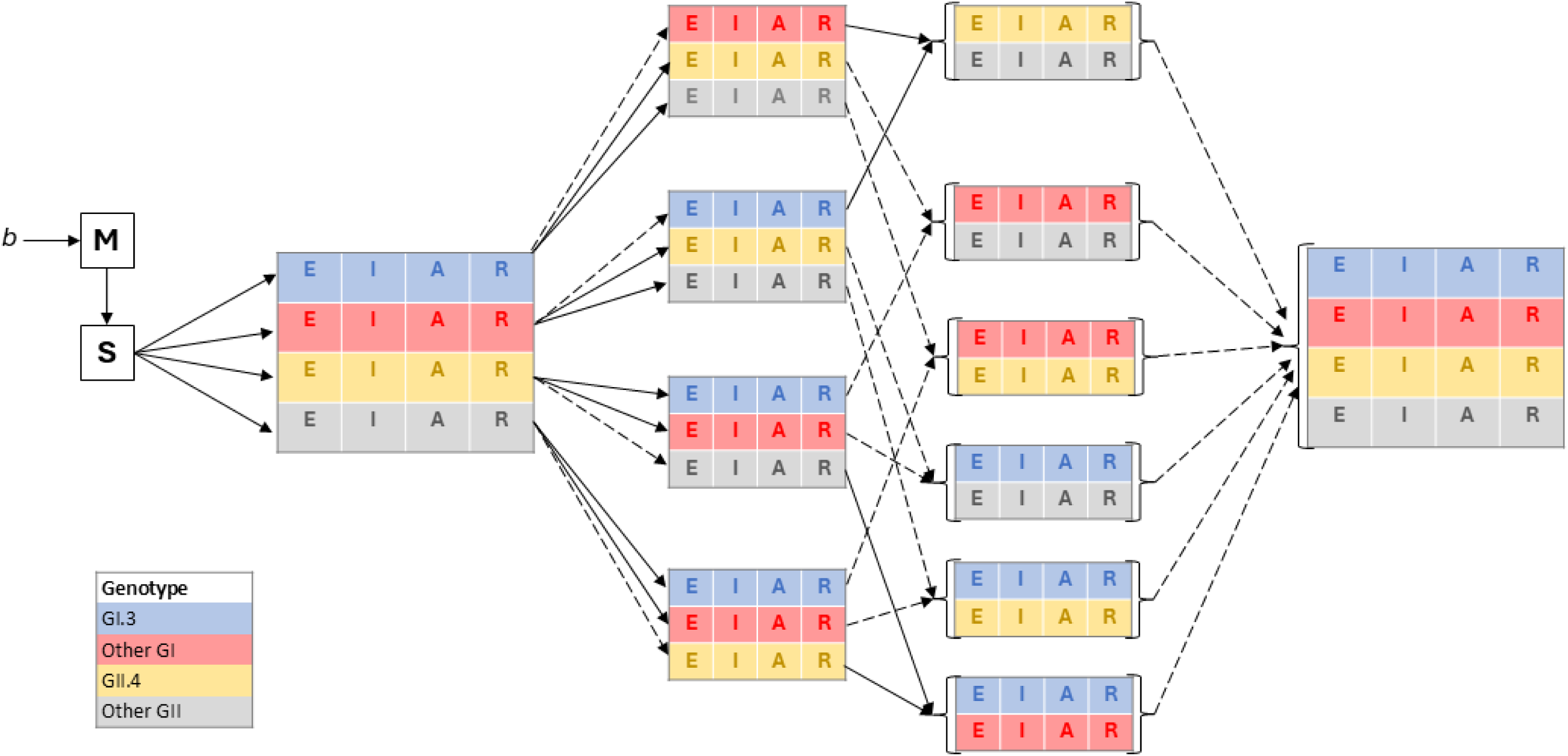
Model structure. Susceptible individuals (S) become infected at a genotype-specific force of infection, progressing to an incubation phase (E) before developing acute gastroenteritis (AGE) (I). Following the symptomatic stage, individuals transition to an asymptomatic viral shedding phase (A) before fully recovering (R). Recovered individuals may acquire symptomatic infection from a different genotype or asymptomatic reinfection from the same genotype. Immunity expands with each symptomatic episode, conferring genotype-specific and cross-genogroup protection (dashed arrows). Waning immunity returns individuals to susceptibility (S) (see Figure 2), though this transition is omitted in the diagram for simplicity. Maternal immunity (M) provides temporary protection for newborns, lasting up to 12 months, after which they transition to susceptibility (S). Newborns without maternal immunity enter the model directly as susceptible individuals.

In our simulation we use contact matrices from the Polymod study (Prem et al., 2017) to capture the average mixing patterns by specific age groups. To account for a more detailed specification of mixing patterns in under 1-year olds we correct our matrices using estimates from van Hoek et al (van Hoek et al., 2013). Our mixing pattern also considers contact changes caused by school term and holidays. During the 2019 coronavirus pandemic (Covid-19), we adjusted the mixing patterns in the model to account for the restrictions on social contacts. Specifically, we incorporated mixing matrices from the CoMix project to reflect these changes (Gimma et al., 2022). **Table 1** describes the fixed model parameters and their sources.

**Table 1.** Model parameters, data sources, parametrisation and mechanistic link to Norovirus epidemiology.

| Fixed model parameters |  |  |  |  |
| --- | --- | --- | --- | --- |
| Parameter | Description |  | Value | Source |
| $\varepsilon$ | Duration of incubation (days) | | 1 | (Devasia et al., 2015) |
| $\theta_{i=1..5}$ | Duration of symptomatic virus shedding in < 5 yro (days) | | 2.5 | (Harris et al., 2019) |
| $\theta_{i>5}$ | Duration of symptomatic virus shedding in > 5 yro (days) | | 1.5 | (Devasia et al., 2015; Harris et al., 2019) |
| $\eta$ | Duration of asymptomatic virus shedding (days) | | 15 | (Atmar et al., n.d.) |
| $\psi$ | Relativeness infectiousness during incubation and asymptomatic period | | 5% | (Simmons et al., n.d.) |
| $\xi$ | Proportion of births assumed to be resistant to GII.4 infection | | 20% | (Lindesmith et al., 2008) |
| $\mu_a$ | Age-specific mortality rate in England | | See reference for values | (Life expectancies - ONS) |
| $c_{ij}$ | Age-specific contact matrix for England | | See reference for values | (Prem et al., 2017b) |
| $cm_{ij}$ | Age-specific contact matrices during COVID-19 in England | | See reference for values | (Gimma et al., 2022) |
| $\omega$ | Wave amplitude of the forced seasonality function | | 0.15 | Manually fitted |
| Calibrated parameters |  |  |  |  |
| Parameter | Description | Posterior (CrI 95%) | Target data | Source of target data |
| $\beta_{1..4}$ | Transmission probability per effective contact | 0.08 (0.06-0.09)<br>0.08 (0.06-0.09)<br>0.14 (0.11-0.16)<br>0.21 (0.16-0.24) | Community Incidence<sup>*</sup><br>Reported time series<sup>‡</sup><br>GII.4 children seroprevalence <sup>±</sup> | (Harris et al., 2019, 2017) (National norovirus and rotavirus report, 27 October 2024) (Lindesmith et al., 2023) |
| $\alpha$ | Relative infectiousness in adults vs children (<15) | 0.67 (0.55-0.86) | Community Incidence<sup>*</sup><br>GII.4 children seroprevalence <sup>±</sup> | (Harris et al., 2019, 2017) (Lindesmith et al., 2023) |
| $\tau$ | Average immunity waning time (in years) | 20.4 (14-29) | Community Incidence<sup>*</sup><br>GII.4 children seroprevalence <sup>±</sup> | (Harris et al., 2019, 2017) (Lindesmith et al., 2023) |
| $\rho_{1..3}$ | Waning immunity factor for secondary infections | 1.7 (1.2-2.1) | Community Incidence<sup>*</sup><br>GII.4 children seroprevalence <sup>±</sup> | (Harris et al., 2019, 2017) (Lindesmith et al., 2023) |
| $\delta_{ab}$ | Average duration of maternal neutralizing AB | 592 (519-672) | GII.4 children seroprevalence <sup>±</sup> | (Lindesmith et al., 2023) |
| $\sigma_1$ | Age specific ratio of community cases to reported cases (0-4) | 110 (94-126) | Reported time series<sup>‡</sup> | (National norovirus and rotavirus report, 27 October 2024) |
| $\sigma_2$ | Age specific ratio of community cases to reported cases (5-15) | 889 (708-1063) | Reported time series<sup>#</sup> | (National norovirus and rotavirus report, 27 October 2024) |
| $\sigma_3$ | Age specific ratio of community cases to reported cases (16-65) | 389 (313-480) | Reported time series<sup>#</sup> | (National norovirus and rotavirus report, 27 October 2024) |
| $\sigma_4$ | Age specific ratio of community cases to reported cases (65+) | 28 (22-35) | Reported time series<sup>#</sup> | (National norovirus and rotavirus report, 27 October 2024) |
| $\Theta^{gi}$ | Cross-protection from GII.4 to other GII | 0.04 (0.02-0.057) | Community Incidence<sup>*</sup><br>GII.4 children seroprevalence<sup>±</sup> | (Lindesmith et al., 2023)<br>(Harris et al., 2019, 2017) |
| $\Theta^{gii}$ | Cross-protection from other GII to GII.4 | 0.013 (0.01-0.015) | Community Incidence<sup>*</sup><br>GII.4 children seroprevalence<sup>±</sup> | (Lindesmith et al., 2023)<br>(Harris et al., 2019, 2017) |

All code was written using R language (see code availability). The model was designed, numerically solved and calibrated using the odin, dust and mcstate R packages (Lees et al., 2021).

### 2.3. Data sources

We calibrate our model against different data sources, that captures three important features of the epidemiology of norovirus viruses:

1. *Norovirus circulation in the community*: The second study of Infectious Intestinal Disease in the community (IID2 study) (Tam et al., 2012) is the largest prospective community cohort in the UK looking at pathogen-specific incidence of gastroenteritis. In this cohort, 6836 healthy participants were followed weekly for 52 weeks and screened for symptoms of acute gastroenteritis. Stool samples from those meeting the case-definition were processed for multiple pathogens, including real-time PCR for norovirus. The age stratified community incidence estimated by Tam et al (Tam et al., 2012) in IID2 was further re-assessed by Harris et al (Harris et al., 2017) to correct for the stringent cycle threshold (ct) used in the original study. We use the latter [26] to build age-specific incidence estimates of Norovirus in the community that we use as target data to compare against our model (**more details in appendix**).
2. *Surveillance and seasonality:* This domain reflects the data as routinely reported by UK Health Security Agency (UKHSA) (4). UKHSA collects, analyses and routinely reports norovirus cases for each season (e.g., July 2024-June 2025), from four sources, namely, the Second-Generation Surveillance System (SGSS), Hospital Norovirus Outbreak Reporting System (HNORS), web-based case and outbreak management system CIMS (Case and Incident Management System), and molecular surveillance produced by UKHSA’s Enteric Virus Unit. In this study we use the first stream (i.e., SGSS) and the fourth stream (molecular surveillance) as it reflects the trends in positive laboratory reports in England and frequency of common genotypes. Importantly, reported cases show a bias towards the 65 year olds and over. This is because outbreaks of acute gastroenteritis are more frequently reported from care-homes and hospital wards. For this reason it is not appropriate to equate these data to estimates of norovirus burden in the community, but instead as a marker of seasonality, and a surrogate indicator of strain distribution.
3. *GII.4 Seroprevalence:* Lindesmith and colleagues obtained sera from 686 young children in the England (1 to 6 years old) from 2008 to 2012. The samples were selected at random from the residual serum archive as explained elsewhere (Lindesmith et al., 2023). Surrogate neutralisation antibody assays were carried out by exposing the serum samples to virus-like particles (VLPs) which reflect the spectrum of emerging pandemic variants of GII.4 from 2005 until 2012 (i.e., Sydney variant). We use this study’s estimates of GII.4 seroprevalence by year of age in children, by aggregating the reported estimates from 2008 to 2012, and compare against model estimates of the same output.

### 2.4. Model calibration

We use an adaptive MCMC within a Bayesian framework to systematically compare our model output to data. Detailed calibration procedures and selected likelihood functions for each data stream can be found in the **supplementary information**.

In **Table 1** we describe the calibrated parameters, and their role in capturing the different target data domains discussed in the ‘Data sources’ section above.

For computational efficiency and to facilitate analysis, we first calibrate the model using a deterministic framework. We then use a stochastic version of the model for subsequent analysis and projections, capturing the inherent randomness of transmission and incorporating parameter estimates from the posterior distribution of the calibrated parameters.

### 2.5. Model selection

The circulation of several simultaneous Norovirus strains imply that a sequence of Norovirus disease events is not only possible but likely over a person’s lifetime. Three central assumptions in our model structure play a central role in this complex process, namely, the potential for asymptomatic reinfection, the average (strain-specific) waning immunity period, and the cross-immunity protection within genogroups. We construct and calibrate the following model configurations to explore the first two assumptions while cross-protection we considered a fixed feature in all configurations (see **Figure 1** for conventions):

1. Waning immunity controlled by one parameter (i.e. all immune individuals, irrespective of how many previous infections (and of what genotype) return to the susceptible class at the same rate, ρ1= ρ2= ρ3=1)
2. Waning immunity with two parameters (that is, individuals who have immunity to one infection (strain) only return to the susceptible class at rate 1/τ, whereas those who have immunity to more than one infection (or strains) return to the susceptible class at a different rate 1/τρ, where ρ1= ρ2= ρ3)
3. Waning immunity with four parameters (one parameter controlling each strain)
4. Waning immunity by one parameter and no asymptomatic reinfection (ρ1= ρ2= ρ3=1)
5. Waning immunity with two parameters and no asymptomatic reinfection (ρ1= ρ2= ρ3)

We fit each of this model configurations to data and used a deviance information criterion (DIC) to select the best fitting model. We use the method described elsewhere (Spiegelhalter et al., 2002).

### 2.6. Analysis

The following set of analyses were performed using the best fitting model structure:

2.6.1. Immunity and cross-protection: We estimate the mean duration of maternal antibodies, mean waning immunity for first and further symptomatic episodes and the genogroup-specific cross-protection.

2.6.2. R_0_ and seasonal Rs estimation: we estimate a basic reproduction number (R_0_) for each strain following the next generation matrix approach (Diekmann et al., 2010). We also report our estimate for the seasonal effective reproductive number (Rs). (see supplementary information)

2.6.3. Lifetime AGE Norovirus episodes accrued: we tracked AGE Norovirus episodes by age and strain to construct an overall estimate of the lifetime number of Norovirus AGE episodes, assuming no further changes to the epidemiology (e.g. further pandemics).

2.6.4. Emergence of a novel GII.4 variant: we evaluate the potential introduction of a new GII.4 strain by adapting the existing immunity levels against GII.4. Using the same model structure, we transfer all existing GII.4 compartments (i.e., E, I, A, R) into the other-GII strain category, and seed 50 new GII.4 cases in the “E” compartment at the start of the simulation (i.e., January).

We explore scenarios of varying levels of immune escape and transmissibility, namely,

1. 0% cross-immunity between GII.4 and other GII strains, 5% less transmissible
2. Calibrated cross-immunity between GII.4 and other GII strains, 5% less transmissible
3. 0% cross-immunity between GII.4 strains and other GII strains, equally transmissible
4. 0% cross-immunity between GII.4 strains and other GII strains, 10% more transmissible
5. 0% cross-immunity between GII.4 and other GII strains, 25% more transmissible

These scenarios are intended to reflect the trade-offs in viral fitness (transmissibility vs immune scape) that could potentially be observed with an emerging variant.

## 3. RESULTS

### 3.1. Model selection

Our systematic model selection demonstrates that existing Norovirus trends are better explained by a model incorporating a two-tier waning immunity structure. Specifically, subsequent infections (beyond the primary infection) extend the average duration of immunity clearance. This means that immunity gained from any secondary or subsequent infection can be represented by a single waning parameter, which in practice is a factor of the waning immunity parameter τ.

Estimates of the calibrated parameters and their epidemiological interpretations are provided in **Table 1**. A comparison of the model with observed data is shown in **Figure 3**. (additional figures on DIC selection included in the supplementary information).

**Figure 3.**
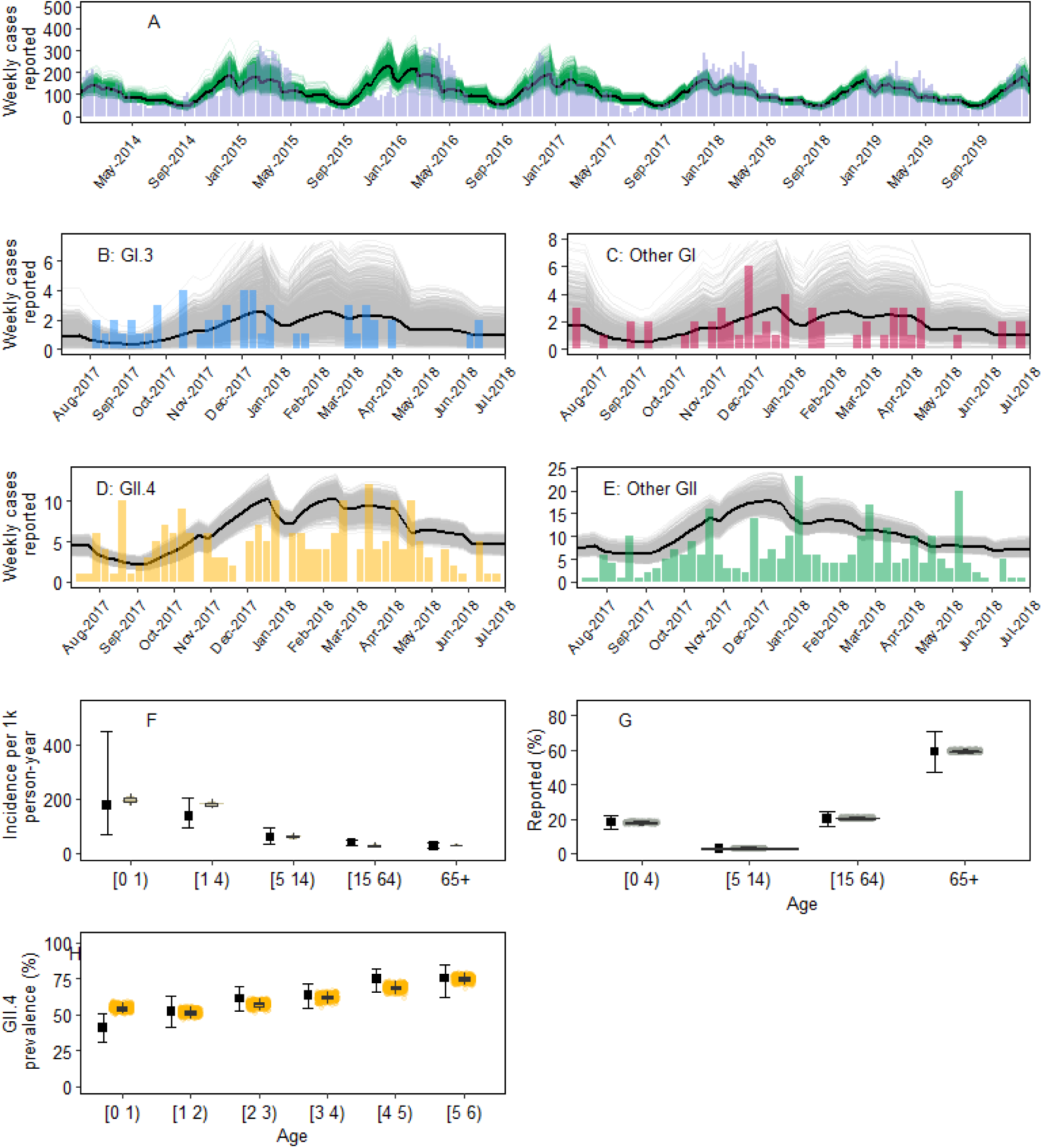
Model calibration to data. A) Reported Norovirus cases in England (grey bar) and 1000 stochastic model trajectories (green lines) and modelled mean (black line); In panels B, C, D and E, modelled reported cases of Norovirus strains (grey shaded area and trajectories) against data on reported frequency of strains (bars) GI.3 (blue), other GI (red), GII.4 (yellow), and other GII (green); F) Box plot of modelled Norovirus incidence of cases per 1000 person-year in the community, by age, against IID2 data (black error bars); G) Mean age distribution of Norovirus reported cases in England as reported to SGSS from 2014 to 2023 (black marker) and modelled age distribution of reported cases (grey violin); H) Seroprevalence of neutralising antibodies against GII.4 genotype among children in England by age (black marker) and modelled GII.4 seroprevalence for the same age groups (yellow violin).

### 3.2. Immunity and cross-protection

In the current model, a mechanism for maternal antibodies protecting against infection during the first months of life is allowed. The current model calibration to data, results in an estimated mean protection of 592 days, after which young children become fully susceptible to infection.

Waning immunity after a first symptomatic episode lasts a mean of 20 years ((i.e., return to full strain-specific susceptibility). Further symptomatic episodes extend the mean duration of protection by a further ∼15 years.

Importantly, given the data, the best fitting mechanism results in estimates of genogroup specific cross-protection (e.g., protection against GII.4 after a single episode of other GII strains) that indicates low cross-protection between GI strains (4%, and negligible protection within the GII group (1%).

### 3.3. R_0_ and Rt estimation

Our basic reproduction number (*R_0_*) estimates show an *R_0_* for GI.3, other GI, GII.4, and other GII, of 2.4 (95% CI 2.2 - 2.7), 2.3 (95% CI 2.1 - 2.5), 3.6 (95% CI 3.4 - 4.1), and 4.4 (95% CI 4.3 – 5.5) respectively **(Figure 4)**. We also estimate a seasonal reproductive number that can be seen in **Figure 4**. The latter account for time variations of contact patterns and the dynamic processes of depletion and replenishment of strain-specific susceptible pools. An important feature of our estimate is the reduction in the GII.4 seasonal reproductive number, which occurs one season after the end of the COVID-19 restrictions. This could be explained, as the dominant transmission strain is replaced temporarily by other-GII strains, which as mentioned above display a higher transmission potential. GII.4 is then expected to re-establish itself as the dominant GII strain after some time (**Figure 4**).

**Figure 4.**
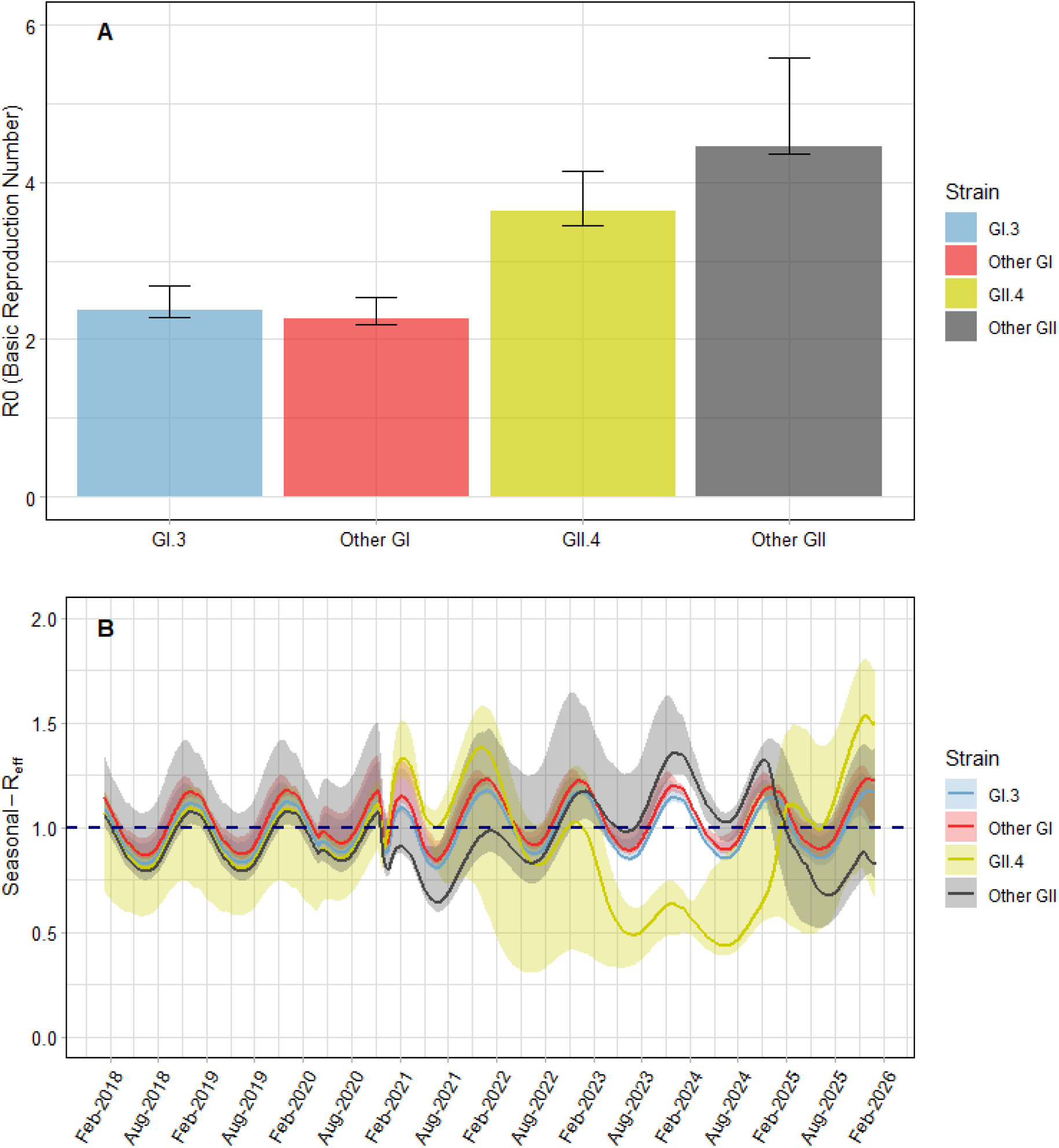
Estimated reproduction number by genotype: A) Estimated R0 by strain – at time 0 assuming a fully susceptible population; B) Seasonal effective reproductive number accounting for seasonal variations and depletion of genotype specific susceptible pools. Solid line shows the mean and shaded area the 95% Credible interval. Dark dashed line marking the R=1 threshold.

### 3.4. Lifetime episodes of disease accrued

By tracking AGE episodes by age, we estimate the number of episodes an average individual will suffer over a lifetime, assuming that infection rates remain the same and no new pandemic strains emerge. We estimate a mean of 3.3 clinical AGE episodes (95% CI 2.8 – 3.6), with the largest contribution being from other GII strains (1.7 95% CI 1.5 – 1.8). Importantly, we estimate that by the age of 4 ∼60% individuals would have suffered at least one symptomatic episode (**Figure 5**).

**Figure 5.**
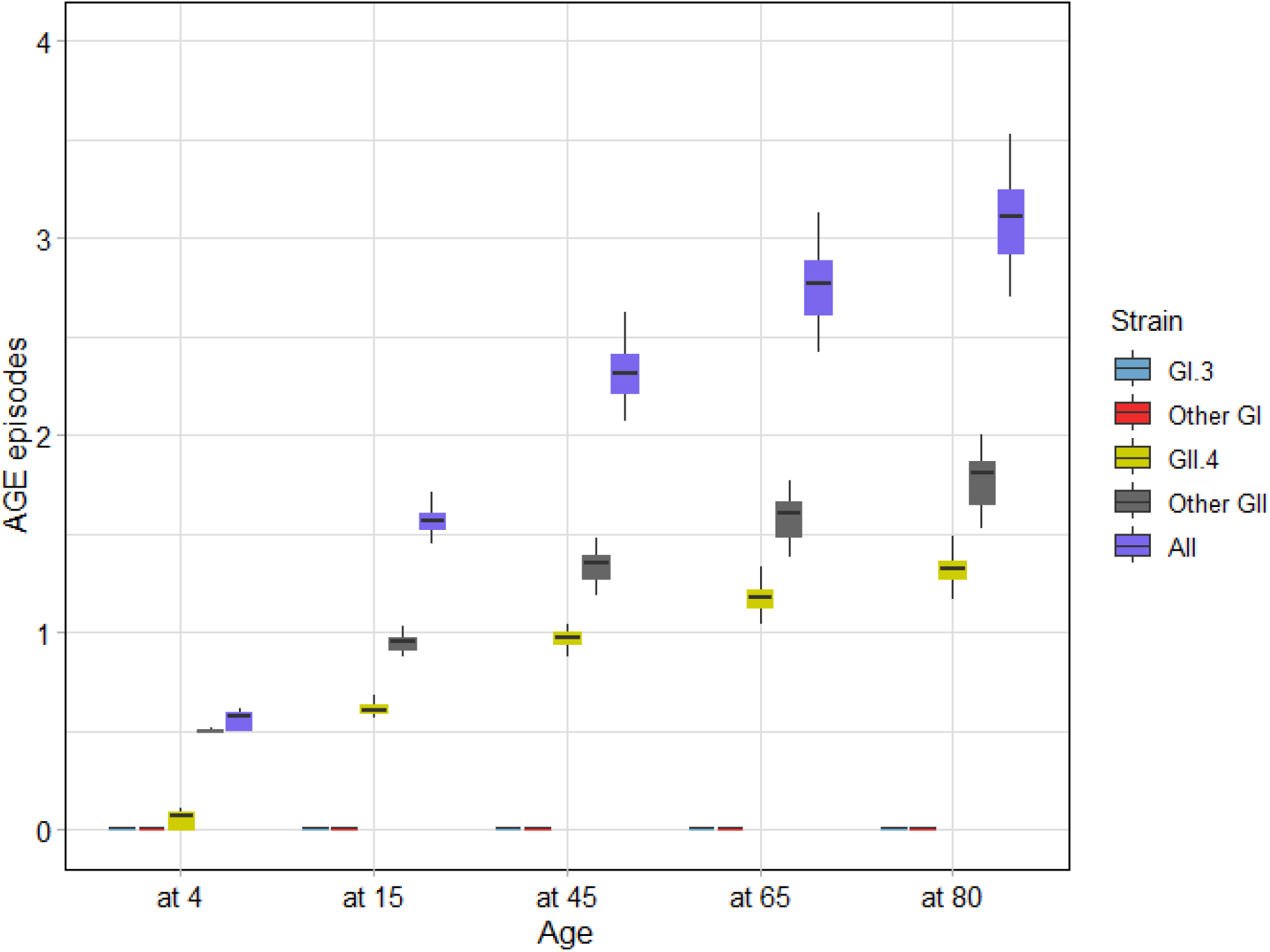
Symptomatic cases over a lifetime: Boxplots showing the modelled number of symptomatic events that an average individual would accrue at different time points over a lifetime assuming rates of infection remain unchanged. E.g., we estimate that an average person would have suffered 1-2 episodes of Norovirus gastroenteritis caused by GII.4 by 80 years old out of a total of 3 or more episodes of disease caused by all genotypes.

### 3.5. Emergence of a novel GII.4 variant

We simulate the introduction of a new GII.4 variant under different assumptions of transmissibility and protection (**Figure 6**, **Table 2).** According to our analysis, an emerging variant which is marginally less transmissible (relative to calibrated transmission levels for GII.4) will result in a collapse of the GII.4 share of AGE cases in the first season after emergence, but will rapidly return to baseline levels, without displaying any significant changes in epidemic size (**Table 2**). An emergent strain with equal transmissibility and no GII cross-protection result in an increase in epidemic size and burden of GII.4 cases in the second season after introduction. Scenarios where the new strain is 10% or 25% more transmissible generate significant increments in epidemic size (up to 56%) from season 1 after introduction as well as an increase in GII.4 burden relative to other strains (**Table 2**). Exploring the age specific burden in these scenarios we see that the baseline pattern (i.e., most cases occurring in the adult population) is not altered by the introduction of a new variant in our simulations (supplementary information Figure S5).

**Figure 6.**
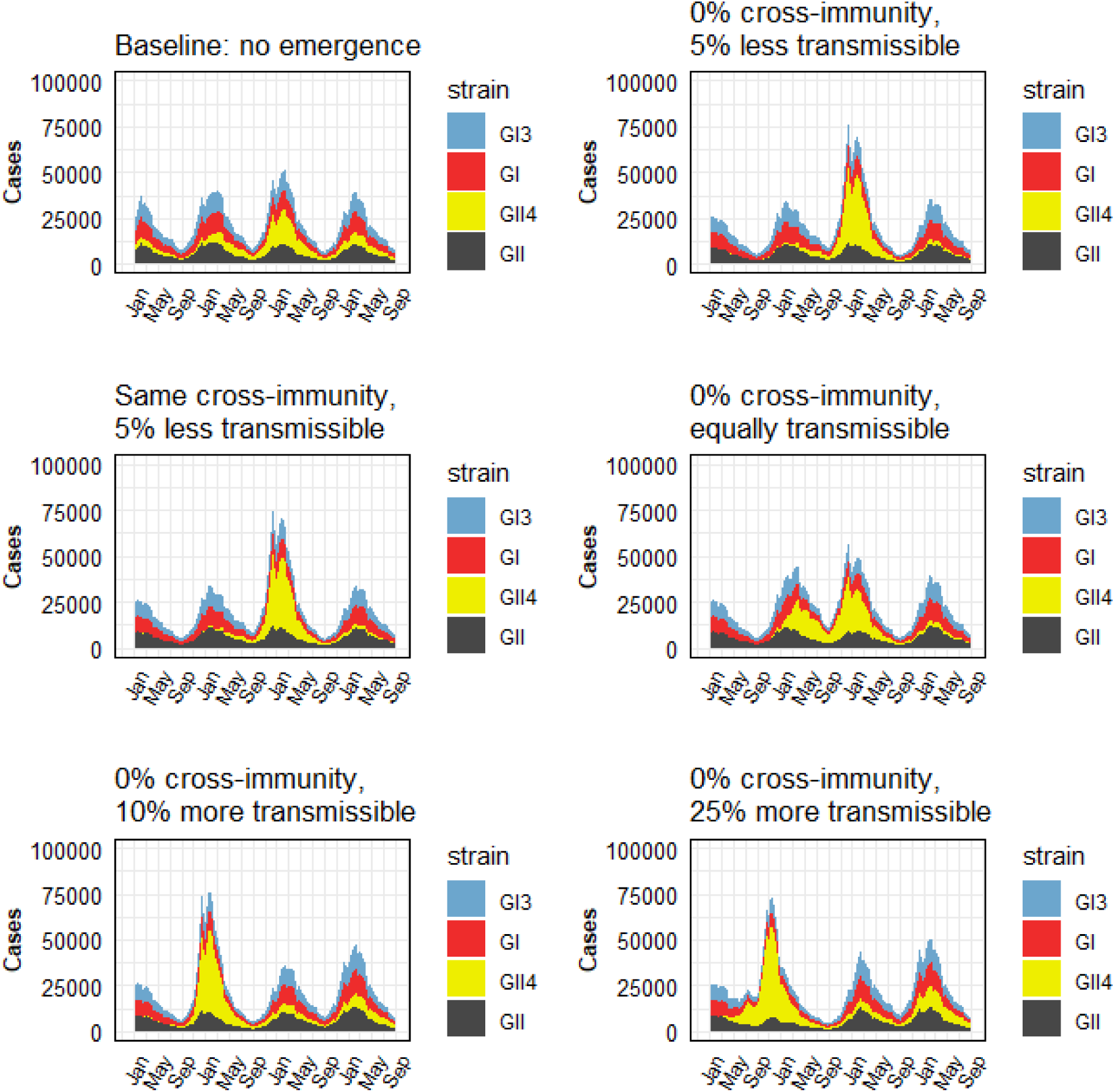
Simulated emergence of a novel GII.4 variant. All panels show the average number of daily modelled Norovirus episodes, stacked by strain, over the simulation time. An emerging GII.4 variant is introduced in January at the beginning of the simulation. Different levels of GII cross-immunity and transmissibility are explored for the new variant.

**Table 2.** Summary of results of emergence of a new GII.4 variant showing scenario characteristics and relative increase in cases by season after introduction.

| Scenario | GII.4 Relative Transmissibility* | GII Cross-protection<sup>‡</sup> | Season 1* |  | Season 2* |  | Season 3* |  |
| --- | --- | --- | --- | --- | --- | --- | --- | --- |
|  |  |  | Relative increase in AGE cases (95% CrI) | Proportion of AGE cases being GII.4 (95% CrI) | Relative increase in AGE cases (95% CrI) | Proportion of AGE cases being GII.4 (95% CrI) | Relative increase in AGE cases (95% CrI) | Proportion of AGE cases being GII.4 |
| Baseline | 100% | 5% (3,6.5%) | 0% (0%,0%) | 31% (12%,52%) | 0% (0%,0%) | 62% (7%,68%) | 0% (0%,0%) | 37% (29%,69%) |
| 1 | 95% | 0% | -65% (-98%,-46%) | 0% (0%,9%) | -1% (-157%,60%) | 59% (0%,75%) | -42% (-171%,51%) | 26% (0%,83%) |
| 2 | 95% | 5% (3,6.5%) | -65% (-100%,-47%) | 0% (0%,6%) | -3% (-151%,57%) | 53% (0%,74%) | -28% (-174%,62%) | 36% (0%,83%) |
| 3 | 100% | 0% | -52% (-88%,-6%) | 4% (0%,37%) | 34% (-147%,68%) | 77% (0%,79%) | -38% (-176%,36%) | 16% (0%,71%) |
| 4 | 110% | 0% | 43% (-51%,67%) | 65% (14%,80%) | 11% (-54%,51%) | 65% (45%,81%) | -12% (-121%,17%) | 28% (13%,37%) |
| 5 | 125% | 0% | 56% (42%,76%) | 80% (76%,87%) | -68% (-99%,-15%) | 36% (30%,46%) | 18% (-43%,34%) | 44% (34%,49%) |

\* Relative to calibrated value of GII.4 transmission probability per effective contact
<sup>‡</sup> Reflects how much protection exists against new GII.4 variant given previous exposure to other-GII strains
\* Epidemic season from July to July in next calendar year.

## 4. DISCUSSION

We developed a mechanistic, age-structured, multi-strain mathematical model to capture the observed trends in Norovirus epidemiology. This model, the first of its kind to be calibrated to multiple England-specific data, integrates both community transmission dynamics and reported data in a Bayesian evidence synthesis. England offers an ideal setting for this analysis due to its range of datasets, including routine surveillance data, community-level active surveillance cohorts, and seroprevalence studies.

Our findings suggest that the average duration of waning immunity is approximately 20 years, with individuals experiencing around 3 to 4 symptomatic episodes over their lifetime (excluding any additional episodes related to new pandemics), which aligns with prior research reporting around 5 episodes per lifetime (Hall et al., 2013). Previous modelling studies have estimated immunity to last around 9 years (Simmons et al., n.d.). This estimate, however, ignores the possibility of re-infection with multiple strains, as it arises from a single-strain transmission model.[35]. Modelling four separate strains will inevitably lead to a longer duration of immunity when compared to a monotypic model that uses the same (or similar) data to fit to. By estimating strain-specific reproduction numbers (R₀), our results confirm the dominance of the GII strains. Previous models that focused on single-strain Norovirus dynamics reported R₀ values between 1.5 and 7 (Gaythorpe et al., 2018b). Our model refines this understanding by highlighting the differential roles of various strains in transmission, thereby offering a more nuanced perspective on reproductive numbers.

Central to our analysis are the estimates of cross-protection between genogroups. For the model structure that best fits the data (i.e., the lowest DIC), cross-protection between GI and GII strains is nearly negligible. This finding has critical implications for vaccine development, particularly for selecting genotypes that can maximize cross-immunity and provide broader protection. Although cross-immunity has been documented to be genogroup specific (Lindesmith et al., 2009; Reeck et al., 2010) its broader impact at the population level remains uncertain (Debbink et al., 2012). Our results underscore the importance of distinguishing between effective protection mechanisms and broader measures of neutralizing antibodies, which may reflect previous exposure but not an active immunity against new infections.

The emergence of new Norovirus variants, particularly within the GII.4 genogroup, remains a significant concern. Recent reports of a novel GII.4 Sydney variant highlight the need to evaluate its potential impact on community-level disease burden (Chhabra et al., 2024a).

Our model predicts that the introduction of an more transmissible emerging novel GII.4 variant could lead to an initial seasonal peak nearly double the average (∼35,000 daily cases). In all scenarios, transmission tends to stabilize after second season.. In England, previous analyses of the emergence of the GII.4 variant Sydney_2012 during the 2012/2013 season revealed an early peak in reported outbreaks. However, this variant did not lead to an overall increase in norovirus activity compared to previous seasons (Allen et al., 2014). Our analysis similarly identifies an early peak in transmission might occur. Scenarios assuming 0% cross-immunity and higher transmissibility align with these findings, as they result in transmission levels consistent with those observed historically.[35]

The model suggests that the interruption of social contact patterns during the COVID-19 pandemic, might lead to a complex epidemiological picture for some years. Our base-case results suggest that GII.4 would predominate during the pandemic, after which it might be replaced by other GII strains, before returning to predominate again. It is interesting to note that GII.17 is currently the most predominant genotype in England and elsewhere.

## 5. LIMITATION AND FURTHER REMARK

Several simplifications were necessary in our model to ensure tractability. For instance, we aggregated less prevalent GI and GII strains, despite evidence that certain strains have public health relevance. Such is the case of the recent surge in GII.17 AGE cases identified in the US and Europe(Chhabra et al., 2024b). Including all possible strains would have introduced considerable complexity without necessarily improving the model’s capacity to answer the motivating questions for our study. Additionally, the waning immunity mechanism in our study does not allow for strain-specific immunity loss, which may better reflect the dynamics of neutralizing antibodies. This limitation arises from the technical challenges of modelling the many possible pathways back to susceptibility in a multi-strain framework.

Although our study does not explicitly simulate the effects of future vaccines, the multi-strain framework we developed paves the way for such analyses. The findings emphasize the critical importance of selecting appropriate genotypes for vaccine formulations and understanding the extent of cross-protection within genogroups to optimize the impact of future immunisation campaigns.

## 6. CONCLUSION

In summary, our study provides critical insights into Norovirus transmission dynamics and immunity, offering an evidence-based foundation for future research and public health interventions. The model we developed is a valuable tool for exploring scenarios involving emerging variants, refining vaccination strategies, and understanding the epidemiological significance of cross-protection and strain-specific dynamics.

## Data Availability

All code and data used for this analysis can be found in the following repository

https://github.com/juanvesga/Norovirus_UK.git

## Acknowledgements

Dr Lisa Lindesmith, Prof Judith Breuer, Dr David Allen, Dr Kathleen O’Reilly of the Noropatrol project for providing feedback during model design.

## CRediT authorship contribution statement

**Juan F. Vesga:** Conceptualization, Methodology, Software, Validation, Formal Analysis, Investigation, Writing – Original Draft, Writing – Review & Editing. **Amy Douglas:** Data Curation, Investigation, Writing – Review & Editing. **Cristina Celma:** Data Curation, Investigation, Writing – Review & Editing. **Edward S. Knock:** Software, Formal Analysis, Writing – Review & Editing. **Marc Baguelin** Conceptualization, Methodology, Software, Writing – Review & Editing. **W. John Edmunds** Conceptualization, Methodology, Supervision, Funding acquisition, Writing – Review & Editing

## Availability of data and materials

All code and data used for this analysis can be found in the following repository https://github.com/juanvesga/Norovirus_UK.git

