## Supplementary information for "The transmission dynamics of Norovirus in England: a genotype-specific modelling study"

### Model Equations

Below the mathematical notation for a deterministic discrete transmission model, used for model calibration, reflecting the best fitted model configuration, that is, the model with 2 immunity parameters and allowing re-infection.

Let:

- $M(t,a)$: Number of individuals carrying GII.4 specific maternal antibodies of age $a$ at time $t$
- $S(t,a)$: Number of susceptible individuals of age $a$ at time $t$
- $E(t,a)$: Number of exposed individuals of age $a$ at time $t$
- $I(t,a)$: Number of symptomatic individuals of age $a$ at time $t$
- $A(t,a)$: Number of asymptomatic individuals of age $a$ at time $t$
- $R(t,a)$: Number of recovered individuals of age $a$ at time $t$

And let superscripts for model states E, I, A and R,

(0): Infection track for naïve individuals

(1): Second Norovirus infection track carrying immunity against strain 1 (i.e., GI.3)

(2): Second Norovirus infection track carrying immunity against strain 2 (i.e., other GI)

(3): Second Norovirus infection track carrying immunity against strain 3 (i.e., GII.4)

(4): Second Norovirus infection track carrying immunity against strain 4 (i.e., other GII)

(12): Third Norovirus infection track carrying immunity against strains 1 and 2 (i.e., GI.3 and other GI)

(13): Third Norovirus infection track carrying immunity against strains 1 and 3 (i.e., GI.3 and GII.4)

(14): Third Norovirus infection track carrying immunity against strains 1 and 4 (i.e., GI.3 and other GII)

(23): Third Norovirus infection track carrying immunity against strains 2 and 3 (i.e., other GI and GII.4)

(24): Third Norovirus infection track carrying immunity against strains 2 and 4 (i.e., other GI and other GII)

(34): Third Norovirus infection track carrying immunity against strains 3 and 4 (i.e., GII.4 and other GII)

(last): Fourth Norovirus infection track carrying immunity against three previous strains except current one

And let $\{1,2,3,4\} \in s$, for strains 1 to 4 , namely, GI.3, other GII, GII.4 and other GII.

The age structure in the model has 14 classes which correspond to the following age ranges: [0 to 1), [1 to 2), [2 to 3), [3 to 4), [4 to 5) , [5 to 6), [6 to 7), [7 to 15), [15 to 25) , [25 to 35), [35 to 45), [45 to 55), [55 to 65), [65 to 75), [75 to 100).

#### Maternal antibodies

$$M(t+1,0) = M(t,0) + B(t)\Omega - M(t,0)\big(\delta + \mu(a)\big) \quad (1)$$

$$M(t+1,a+1) = M(t,a) - M(t,a)\big(\delta + \mu(a)\big), for\ a > 0 \quad (2)$$

#### Susceptible

$$S(t+1,0) = S(t,0) + B(t)(1-\Omega) + M(t,0)\delta - \sum_{a',s} \lambda_s(t,a')\, S(t,0) - S(t,0)\mu(0) \quad (3)$$

$$\begin{gathered} S(t+1,a) = S(t,a) + M(t,a)\delta + R_s^{(0)}(t,a)\tau + \\ \tau\rho \begin{pmatrix} R_s^{(1)}(t,a) + R_s^{(2)}(t,a) + R_s^{(3)}(t,a) + R_s^{(4)}(t,a) + R_s^{(12)}(t,a) + \\ R_s^{(13)}(t,a) + R_s^{(14)}(t,a) + R_s^{(23)}(t,a) + R_s^{(24)}(t,a) + R_s^{(34)}(t,a) + R_s^{(last)}(t,a) \end{pmatrix} \\ - \sum_{a',s} \lambda_s(t,a')\, S(t,a) - S(t,a)\mu(a), \\ for\ a > 0 \end{gathered} \quad (4)$$

#### First infection

$$E_s^{(0)}(t+1,a+1) = E_s^{0}(t,a) + S(t,a)\lambda_s(t,a) - E_s^{0}(t,a)\big(\varepsilon + \mu(a)\big) \quad (5)$$

$$I_s^{(0)}(t+1,a+1) = I_s^{(0)}(t,a) + E_s^{(0)}(t,a)\varepsilon - I_s^{0}(t,a)\big(\theta(a) + \mu(a)\big) \quad (6)$$

$$\begin{gathered} A_s^{(0)}(t+1,a+1) = A_s^{(0)}(t,a) + R_s^{(0)}(t,a)\lambda_{s'}(t,a) + I_s^{(0)}(t,a)\theta(a) - A_s^{(0)}(t,a)\big(\eta + \mu(a)\big), \\ for\ s' = s \end{gathered} \quad (7)$$

$$A_s^{(0)}(t+1,a+1) = A_s^{(0)}(t,a) + I_s^{(0)}(t,a)\theta(a) - A_s^{(0)}(t,a)\big(\eta + \mu(a)\big), \qquad for\ s' \neq s \quad (8)$$

$$R_s^{(0)}(t+1,a+1) = R_s^{(0)}(t,a) + A_s^{(0)}(t,a)\eta - \sum_{s'=1}^{4} R_s^{(0)}(t,a)\lambda_{s'}(t,a) - R_s^{(0)}(t,a)(\mu(a)+\tau) \quad (9)$$

Second infection, after GI.3

$$E_s^{(1)}(t+1,a+1) = E_s^{(1)}(t,a) + R_1^{(0)}(t,a)\lambda_s(t,a) - E_s^{(1)}(t,a)\big(\varepsilon + \mu(a)\big) \quad (10)$$

$$I_s^{(1)}(t+1,a+1) = I_s^{(1)}(t,a) + E_s^{(1)}(t,a)\varepsilon - I_s^{(1)}(t,a)\big(\theta(a) + \mu(a)\big) \quad (11)$$

$$A_s^{(1)}(t+1,a+1) = A_s^{(1)}(t,a) + R_s^{(1)}(t,a)\lambda_{s'}(t,a) + I_s^{(1)}(t,a)\theta(a) - A_s^{(1)}(t,a)\big(\eta + \mu(a)\big), \quad for\ s' = s \quad (12)$$

$$A_s^{(1)}(t+1,a+1) = A_s^{(1)}(t,a) + I_s^{(1)}(t,a)\theta(a) - A_s^{(1)}(t,a)\big(\eta + \mu(a)\big), \qquad for\ s' \neq s \quad (13)$$

$$R_s^{(1)}(t+1,a+1) = R_s^{(1)}(t,a) + A_s^{(1)}(t,a)\eta - \sum_{s' \in (2,3,4)} R_s^{(1)}(t,a)\lambda_{s'}(t,a) - R_s^{(1)}(t,a)(\mu(a)+\tau\rho) \quad (14)$$

Second infection, after other GI

$$E_s^{(2)}(t+1,a+1) = E_s^{(2)}(t,a) + R_2^{(0)}(t,a)\lambda_s(t,a) - E_s^{(2)}(t,a)\big(\varepsilon + \mu(a)\big) \quad (15)$$

$$I_s^{(2)}(t+1,a+1) = I_s^{(2)}(t,a) + E_s^{(2)}(t,a)\varepsilon - I_s^{(2)}(t,a)\big(\theta(a) + \mu(a)\big) \quad (17)$$

$$A_s^{(2)}(t+1,a+1) = A_s^{(2)}(t,a) + R_s^{(2)}(t,a)\lambda_{s'}(t,a) + I_s^{(2)}(t,a)\theta(a) - A_s^{(2)}(t,a)\big(\eta + \mu(a)\big), \quad for\ s' = s \tag{18}$$

$$A_s^{(2)}(t+1,a+1) = A_s^{(2)}(t,a) + I_s^{(2)}(t,a)\theta(a) - A_s^{(2)}(t,a)\big(\eta + \mu(a)\big), \qquad for\ s' \neq s \tag{19}$$

$$R_s^{(2)}(t+1,a+1) = R_s^{(2)}(t,a) + A_s^{(2)}(t,a)\eta - \sum_{s' \in (1,3,4)} R_s^{(2)}(t,a)\lambda_{s'}(t,a) - R_s^{(2)}(t,a)(\mu(a) + \tau\rho) \tag{20}$$

<u>Second infection, after GII.4</u>

$$E_s^{(3)}(t+1,a+1) = E_s^{(3)}(t,a) + R_3^{(0)}(t,a)\lambda_s(t,a) - E_s^{(3)}(t,a)\big(\varepsilon + \mu(a)\big) \tag{21}$$

$$I_s^{(3)}(t+1,a+1) = I_s^{(3)}(t,a) + E_s^{(3)}(t,a)\varepsilon - I_s^{(3)}(t,a)\big(\theta(a) + \mu(a)\big) \tag{22}$$

$$A_s^{(3)}(t+1,a+1) = A_s^{(3)}(t,a) + R_s^{(3)}(t,a)\lambda_{s'}(t,a) + I_s^{(3)}(t,a)\theta(a) - A_s^{(3)}(t,a)\big(\eta + \mu(a)\big), \quad for\ s' = s \tag{23}$$

$$A_s^{(3)}(t+1,a+1) = A_s^{(3)}(t,a) + I_s^{(3)}(t,a)\theta(a) - A_s^{(3)}(t,a)\big(\eta + \mu(a)\big), \qquad for\ s' \neq s \tag{24}$$

$$R_s^{(3)}(t+1,a+1) = R_s^{(3)}(t,a) + A_s^{(3)}(t,a)\eta - \sum_{s' \in (1,2,4)} R_s^{(3)}(t,a)\lambda_{s'}(t,a) - R_s^{(3)}(t,a)(\mu(a) + \tau\rho) \tag{25}$$

Second infection, after other GII

$$E_s^{(4)}(t+1,a+1) = E_s^{(4)}(t,a) + R_4^{(0)}(t,a)\lambda_s(t,a) - E_s^{(4)}(t,a)\big(\varepsilon + \mu(a)\big) \qquad (26)$$

$$I_s^{(4)}(t+1,a+1) = I_s^{(4)}(t,a) + E_s^{(4)}(t,a)\varepsilon - I_s^{4)}(t,a)\big(\theta(a) + \mu(a)\big) \qquad (27)$$

$$A_s^{(4)}(t+1,a+1) = A_s^{(4)}(t,a) + R_s^{(4)}(t,a)\lambda_{s'}(a) + I_s^{(4)}(t,a)\theta(a) - A_s^{(4)}(t,a)\big(\eta + \mu(a)\big),$$
$$for\ s' = s \qquad (28)$$

$$A_s^{(4)}(t+1,a+1) = A_s^{(4)}(t,a) + I_s^{(4)}(t,a)\theta(a) - A_s^{(4)}(t,a)\big(\eta + \mu(a)\big), \qquad for\ s' \neq s \qquad (29)$$

$$R_s^{(4)}(t+1,a+1) = R_s^{(4)}(t,a) + A_s^{(4)}(t,a)\eta$$
$$- \sum_{s' \in (1,2,3)} R_s^{(4)}(t,a)\lambda_{s'}(t,a) - R_s^{(4)}(t,a)(\mu(a) + \tau\rho) \qquad (30)$$

Third infection, after GI.3 and other GI

$$E_s^{(12)}(t+1,a+1) = E_s^{(12)}(t,a) + R_2^{(1)}(t,a)\lambda_s(t,a) + R_1^{(2)}(t,a)\lambda_s(t,a)$$
$$-E_s^{(12)}(t,a)\big(\varepsilon + \mu(a)\big) \qquad (31)$$

$$I_s^{(12)}(t+1,a+1) = I_s^{(12)}(t,a) + E_s^{(12)}(t,a)\varepsilon - I_s^{(12)}(t,a)\big(\theta(a) + \mu(a)\big) \qquad (32)$$

$$A_s^{(12)}(t+1,a+1) = A_s^{(12)}(t,a) + R_s^{(12)}(t,a)\lambda_{s'}(a) + I_s^{(12)}(t,a)\theta(a) - A_s^{(12)}(t,a)\big(\eta + \mu(a)\big),$$
$$for\ s' = s \qquad (33)$$

$$A_s^{(12)}(t+1,a+1) = A_s^{(12)}(t,a) + I_s^{(12)}(t,a)\theta(a) - A_s^{(12)}(t,a)\big(\eta + \mu(a)\big), \qquad for\ s' \neq s \quad (34)$$

$$R_s^{(12)}(t+1,a+1) = R_s^{(12)}(t,a) + A_s^{(12)}(t,a)\eta$$
$$- \sum_{s' \in (3,4)} R_s^{(12)}(t,a)\lambda_{s'}(t,a) - R_s^{(12)}(t,a)(\mu(a) + \tau\rho) \quad (35)$$

Third infection, after GI.3 and GII.4

$$E_s^{(13)}(t+1,a+1) = E_s^{(13)}(t,a) + R_3^{(1)}(t,a)\lambda_s(t,a) + R_1^{(3)}(t,a)\lambda_s(t,a)$$
$$-E_s^{(13)}(t,a)\big(\varepsilon + \mu(a)\big) \quad (36)$$

$$I_s^{(13)}(t+1,a+1) = I_s^{(13)}(t,a) + E_s^{(13)}(t,a)\varepsilon - I_s^{(13)}(t,a)\big(\theta(a) + \mu(a)\big) \quad (37)$$

$$A_s^{(13)}(t+1,a+1) = A_s^{(13)}(t,a) + R_s^{(13)}(t,a)\lambda_{s'}(a) + I_s^{(13)}(t,a)\theta(a) - A_s^{(13)}(t,a)\big(\eta + \mu(a)\big),$$
$$for\ s' = s \quad (38)$$

$$A_s^{(13)}(t+1,a+1) = A_s^{(13)}(t,a) + I_s^{(13)}(t,a)\theta(a) - A_s^{(13)}(t,a)\big(\eta + \mu(a)\big),$$
$$for\ s' \neq s \quad (39)$$

$$R_s^{(13)}(t+1,a+1) = R_s^{(13)}(t,a) + A_s^{(13)}(t,a)\eta$$
$$- \sum_{s' \in (1,3)} R_s^{(13)}(t,a)\lambda_{s'}(t,a) - R_s^{(13)}(t,a)(\mu(a) + \tau\rho) \quad (40)$$

Third infection, after GI.3 and other GII

$$E_s^{(14)}(t+1,a+1) = E_s^{(14)}(t,a) + R_4^{(1)}(t,a)\lambda_s(t,a) + R_1^{(4)}(t,a)\lambda_s(t,a) - E_s^{(14)}(t,a)\big(\varepsilon + \mu(a)\big) \quad (41)$$

$$I_s^{(14)}(t+1,a+1) = I_s^{(14)}(t,a) + E_s^{(14)}(t,a)\varepsilon - I_s^{(14)}(t,a)\big(\theta(a) + \mu(a)\big) \quad (42)$$

$$A_s^{(14)}(t+1,a+1) = A_s^{(14)}(t,a) + R_s^{(14)}(t,a)\lambda_{s'}(a) + I_s^{(14)}(t,a)\theta(a) - A_s^{(14)}(t,a)\big(\eta + \mu(a)\big), \quad for\ s' = s \quad (43)$$

$$A_s^{(14)}(t+1,a+1) = A_s^{(14)}(t,a) + I_s^{(14)}(t,a)\theta(a) - A_s^{(14)}(t,a)\big(\eta + \mu(a)\big), \quad for\ s' \neq s \quad (44)$$

$$R_s^{(14)}(t+1,a+1) = R_s^{(14)}(t,a) + A_s^{(14)}(t,a)\eta - \sum_{s' \in (3,4)} R_s^{(14)}(t,a)\lambda_{s'}(t,a) - R_s^{(14)}(t,a)(\mu(a) + \tau\rho) \quad (45)$$

<u>Third infection, after other GII and GII.4</u>

$$E_s^{(23)}(t+1,a+1) = E_s^{(23)}(t,a) + R_3^{(2)}(t,a)\lambda_s(t,a) + R_2^{(3)}(t,a)\lambda_s(t,a) - E_s^{(23)}(t,a)\big(\varepsilon + \mu(a)\big) \quad (46)$$

$$I_s^{(23)}(t+1,a+1) = I_s^{(23)}(t,a) + E_s^{(23)}(t,a)\varepsilon - I_s^{(23)}(t,a)\big(\theta(a) + \mu(a)\big) \quad (47)$$

$$A_s^{(23)}(t+1,a+1) = A_s^{(23)}(t,a) + R_s^{(23)}(t,a)\lambda_{s'}(a) + I_s^{(23)}(t,a)\theta(a) - A_s^{(23)}(t,a)\big(\eta + \mu(a)\big), \quad for\ s' = s \quad (48)$$

$$A_s^{(23)}(t+1,a+1) = A_s^{(23)}(t,a) + \; I_s^{(23)}(t,a)\theta(a) - A_s^{(23)}(t,a)\big(\eta + \mu(a)\big),$$
$$for\; s' \neq s \tag{49}$$

$$R_s^{(23)}(t+1,a+1) = R_s^{(23)}(t,a) + \; A_s^{(23)}(t,a)\eta$$
$$- \sum_{s' \in (1,4)} R_s^{(23)}(t,a)\lambda_{s'}(t,a) - R_s^{(23)}(t,a)(\mu(a) + \tau\rho) \tag{50}$$

<u>Third infection, after other GII and other GII</u>

$$E_s^{(24)}(t+1,a+1) = E_s^{(24)}(t,a) + \; R_4^{(2)}(t,a)\lambda_s(t,a) + R_2^{(4)}(t,a)\lambda_s(t,a)$$
$$-E_s^{(24)}(t,a)\big(\varepsilon + \mu(a)\big) \tag{51}$$

$$I_s^{(24)}(t+1,a+1) = I_s^{(24)}(t,a) + \; E_s^{(24)}(t,a)\varepsilon - I_s^{(24)}(t,a)\big(\theta(a) + \mu(a)\big) \tag{52}$$

$$A_s^{(24)}(t+1,a+1) = A_s^{(24)}(t,a) + \; R_s^{(24)}(t,a)\lambda_{s'}(a) + \; I_s^{(24)}(t,a)\theta(a) - A_s^{(24)}(t,a)\big(\eta + \mu(a)\big),$$
$$for\; s' = s \tag{53}$$

$$A_s^{(24)}(t+1,a+1) = A_s^{(24)}(t,a) + \; I_s^{(24)}(t,a)\theta(a) - A_s^{(24)}(t,a)\big(\eta + \mu(a)\big),$$
$$for\; s' \neq s \tag{54}$$

$$R_s^{(24)}(t+1,a+1) = R_s^{(24)}(t,a) + \; A_s^{(24)}(t,a)\eta$$
$$- \sum_{s' \in (1,3)} R_s^{(24)}(t,a)\lambda_{s'}(t,a) - R_s^{(24)}(t,a)(\mu(a) + \tau\rho) \tag{55}$$

Third infection, after GII4 and other GII

$$E_s^{(34)}(t+1,a+1) = E_s^{(34)}(t,a) + R_4^{(3)}(t,a)\lambda_s(t,a) + R_3^{(4)}(t,a)\lambda_s(t,a) \\ -E_s^{(34)}(t,a)\big(\varepsilon + \mu(a)\big) \quad (56)$$

$$I_s^{(34)}(t+1,a+1) = I_s^{(34)}(t,a) + E_s^{(34)}(t,a)\varepsilon - I_s^{(34)}(t,a)\big(\theta(a) + \mu(a)\big) \quad (56)$$

$$A_s^{(34)}(t+1,a+1) = A_s^{(34)}(t,a) + R_s^{(34)}(t,a)\lambda_{s'}(a) + I_s^{(34)}(t,a)\theta(a) - A_s^{(34)}(t,a)\big(\eta + \mu(a)\big), \\ for\ s' = s \quad (57)$$

$$A_s^{(34)}(t+1,a+1) = A_s^{(34)}(t,a) + I_s^{(34)}(t,a)\theta(a) - A_s^{(34)}(t,a)\big(\eta + \mu(a)\big), \\ for\ s' \neq s \quad (58)$$

$$R_s^{(34)}(t+1,a+1) = R_s^{(34)}(t,a) + A_s^{(34)}(t,a)\eta \\ - \sum_{s' \in (1,2)} R_s^{(34)}(t,a)\lambda_{s'}(t,a) - R_s^{(34)}(t,a)(\mu(a) + \tau\rho) \quad (59)$$

Fourth infection

$$E_s^{(last)}(t+1,a+1) = \begin{cases} E_1^{(last)}(t,a) + (R_4^{(23)}(t,a) + R_3^{(24)}(t,a) + R_2^{(34)}(t,a))\lambda_1(t,a) \\ \quad -E_1^{(last)}(t,a)\big(\varepsilon + \mu(a)\big) \\ E_2^{(last)}(t,a) + (R_4^{(13)}(t,a) + R_3^{(14)}(t,a) + R_1^{(34)}(t,a))\lambda_2(t,a) \\ \quad -E_2^{(last)}(t,a)\big(\varepsilon + \mu(a)\big) \\ E_3^{(last)}(t,a) + (R_4^{(12)}(t,a) + R_2^{(14)}(t,a) + R_1^{(24)}(t,a))\lambda_4(t,a) \\ \quad -E_3^{(last)}(t,a)\big(\varepsilon + \mu(a)\big) \\ E_4^{(last)}(t,a) + (R_3^{(12)}(t,a) + R_2^{(13)}(t,a) + R_1^{(23)}(t,a))\lambda_4(t,a) \\ \quad -E_4^{(last)}(t,a)\big(\varepsilon + \mu(a)\big) \end{cases} \quad (60)$$

$$I_s^{(last)}(t+1,a+1) = I_s^{(last)}(t,a) + E_s^{(last)}(t,a)\varepsilon - I_s^{(last)}(t,a)\big(\theta(a)+\mu(a)\big) \quad (61)$$

$$A_s^{(last)}(t+1,a+1) = A_s^{(last)}(t,a) + R_s^{(last)}(t,a)\lambda_{s'}(a) + I_s^{(last)}(t,a)\theta(a) - A_s^{(last)}(t,a)\big(\eta+\mu(a)\big), \quad for\ s' = s \quad (62)$$

$$A_s^{(last)}(t+1,a+1) = A_s^{(last)}(t,a) + I_s^{(34)}(t,a)\theta(a) - A_s^{(last)}(t,a)\big(\eta+\mu(a)\big), \quad for\ s' \neq s \ (63)$$

$$R_s^{(last)}(t+1,a+1) = R_s^{(last)}(t,a) + A_s^{(last)}(t,a)\eta - \sum_{s'=s} R_s^{(last)}(t,a)\lambda_{s'}(t,a) - R_s^{(last)}(t,a)(\mu(a)+\tau\rho) \quad (65)$$

Force of infection

$$\lambda_s(t,a) = \beta_s(t)\sum_{g=1}^{G}\sum_{a'=1}^{A} c_{a,a'}\frac{\Big(E_s^{(g)}(t,a')\psi + I_s^{(g)}(t,a') + A_s^{(g)}(t,a')\psi\Big)}{N}\alpha(a') \quad (66)$$

Where A is the total age categories and G contains all the superscripts for infection track, and $\beta_s(t)$ is the seasonally varying transmission rate. To account for seasonality, the transmission rate $\beta_s(t)$ is modelled as:

$$\beta_s(t) = \beta_s(1 + \alpha\cos(2\pi t/T)) \quad (67)$$

where:

- $\beta_s$ is the baseline transmission rate,
- $\alpha$ is the amplitude of seasonal variation,
- $T$ is the period of seasonality (e.g., 1 year).
- $C_{a,a'}$ is the symmetric contact matrix for individuals of age a with those age a'

For the model projections beyond the calibrated targets we switch the above model to a stochastic form, in which we change the mean transition rate multiplication for a binomial draw.

### Model calibration

We denote by θ the vector of input parameters, for all model inputs subject to uncertainty. For a given parameter set θ, we followed the steps:

1. Run an instance of the deterministic model
2. Record model output:
    a. Cross-sectional GII.4 prevalence by age in children 0 to 7
    b. Reported Norovirus cases over a period 2014 to 2019
    c. Age and strain specific community incidence at a point in time matching that of data source (IID2 ~208/2009)
    d. Age distribution of reported cases
3. Calculate global posterior for the model and calibration targets described in table 1.

To compare these model projections with data D, we defined the posterior density π(θ) as:

$$\Pi(\theta) \propto \mathrm{L}(\mathrm{D}|\theta).\,\mathrm{P}(\theta) \qquad (68)$$

Where L is the likelihood of the data D given models with parameters θ , and P is the joint prior distribution for θ. For P, we took independent distributions. The likelihood L was constructed as follows. For count outputs (i.e., reported cases) we estimate the negative binomial likelihood, and the binomial likelihood for binary outcomes like prevalence. For a given parameter set θ, we then constructed the overall likelihood π(θ) as a product of these distributions over all calibration targets listed in table 1 and S1. In practice we computed the logarithm of π(θ), thus taking the sum of the logarithms of each of the probability densities involved.

With π(θ) thus defined, we sampled the posterior density using a Markov Chain Monte Carlo approach. In brief, this approach implements a random walk through the space of parameter values θ to obtain an unbiased sample of the posterior density. We implemented the Metropolis Hastings algorithm with adaptive gaussian proposal existing in the package “Dust” for Odin models in R. For the set of parameter values thus obtained, we took every tenth element to reduce autocorrelation, thus yielding an ‘ensemble’ of parameters θ1,θ2,… ; This ensemble captures simultaneously the uncertainty in the parameter inputs, as well as in the calibration data. Then, to estimate uncertainty in a given simulated output φ (e.g. in the projected incidence of GII.4), we simulated this output φi for every θi. We finally estimated uncertainty in φi by determining its 2.5th, 50th and 97.5th percentiles. For graphic depiction of model fits to data see figure 3 in the main text. **Figure S1** shows the MCMC trace plots for the posterior value showing mixing, and **Figure S2-S3** shows density, histograms and violin plots for the calibrated parameters.

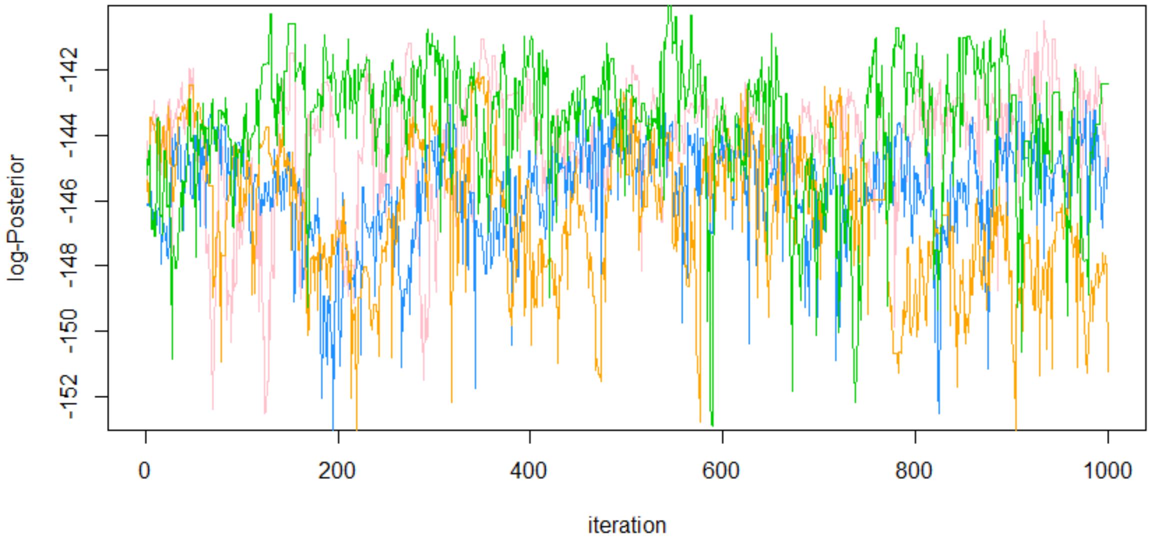

Figure S1: Trace plots of four chains showing log-posteriors

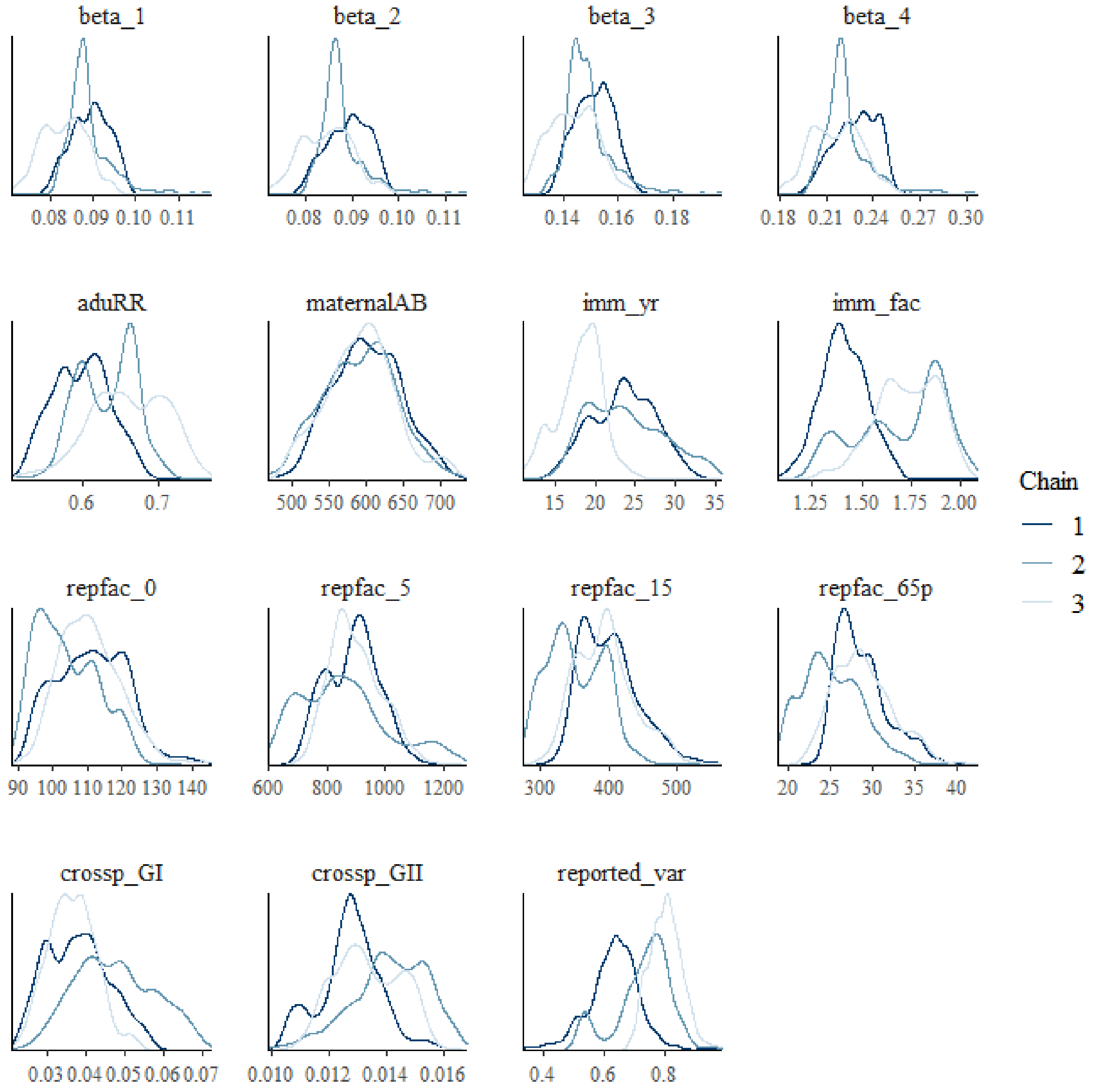

Figure S2: Density plots of calibrated parameters by chain

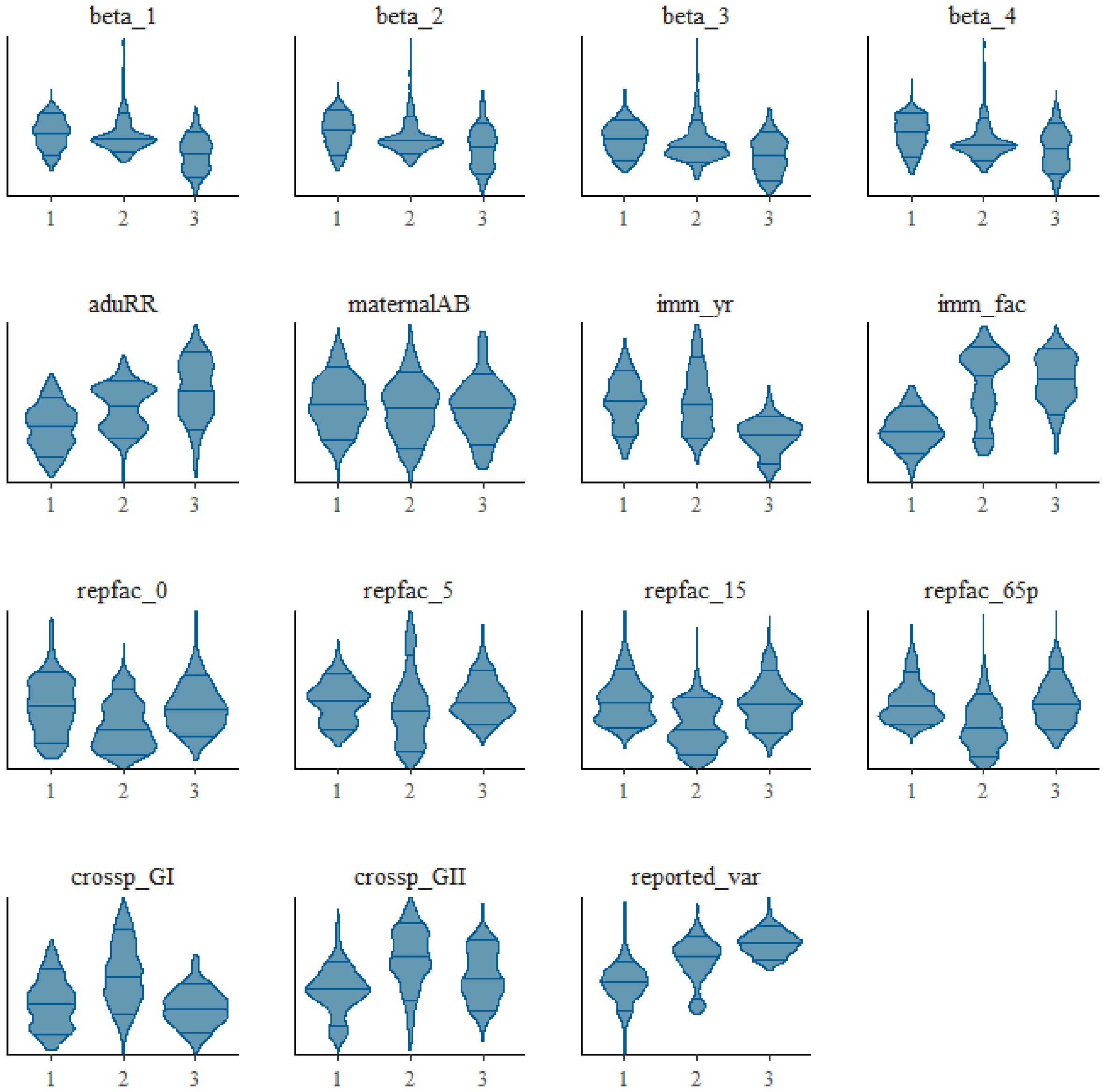

Figure S3: Density violin plots of calibrated parameters by chain

Figure S4: MCM Trace plots of calibrated parameters by chain

Convergence was assessed visually by inspecting the trace plots of the calibrated parameters and also through estimation of the Gelman –Rubin convergence diagnostic, computed as follows:

$$\ddot{R} = \frac{\ddot{V}}{\mathrm{w}} \tag{69}$$

Where v¨ is the posterior variance estimate of the combined chains and w is the within-chain variance. If the chains have converged to the target posterior distribution, then R¨ (also known as the scale reduction factor) should be close to 1. As a rule of thumb, values below 1.1 are typically considered to indicate convergence.

### DIC and model selection

We assess structural assumptions on immunity and reinfection by performing a model selection procedure. We use the Deviance information criterion (DIC). For this we fit each model configuration (see figure 2 in main text for details),

1) Waning immunity controlled by one parameter (i.e. all immune individuals, irrespective of how many previous infections (and of what genotype) return to the susceptible class at the same rate, $\rho 1 = \rho 2 = \rho 3 = 1$)
2) Waning immunity with two parameters (that is, individuals who have immunity to one infection (strain) only return to the susceptible class at rate $1/\tau$, whereas those who have immunity to more than one infection (or strains) return to the susceptible class at a different rate $1/\tau\rho$, where $\rho 1 = \rho 2 = \rho 3$)
3) Waning immunity with four parameters (one parameter controlling each strain)
4) Waning immunity by one parameter and no asymptomatic reinfection ($\rho 1 = \rho 2 = \rho 3 = 1$)
5) Waning immunity with two parameters and no asymptomatic reinfection ($\rho 1 = \rho 2 = \rho 3$)

For each model we estimate the DIC as follows:

We first define the deviance $D(\theta)$ as a measure of model fit and given:

$$\mathrm{D}(\theta) = -2 \log \mathrm{p}(\mathrm{y} \mid \theta) \tag{70}$$

Where $y$ is the observed data, $\theta$ represents the model parameters, $p(y|\theta)$ is the likelihood of the data given the parameters.

We then compute the posterior mean deviance $\bar{D}$, which is the average deviance over the posterior distribution of the parameters, and measures the average fit of the model to the data.

$$\bar{\mathrm{D}} = \mathrm{E}\theta \mid \mathrm{y}[\mathrm{D}(\theta)] \tag{71}$$

To capture the complexity of each model we estimate the effective number of parameters $pD$ captures the complexity of the model and is computed as:

$$pD = \bar{D} - D(\bar{\theta}) \tag{72}$$

Where $D(\bar{\theta})$ is the deviance evaluated at the posterior mean of the parameters $\bar{\theta}$.

And finally estimate DIC as:

$$\mathrm{DIC} = \bar{D} + pD \tag{73}$$

Finally, to select the best model given the assumptions, model fit and model complexity we choose the lowest values of DIC. **Figure S4** shows the DIC for the different model configurations.

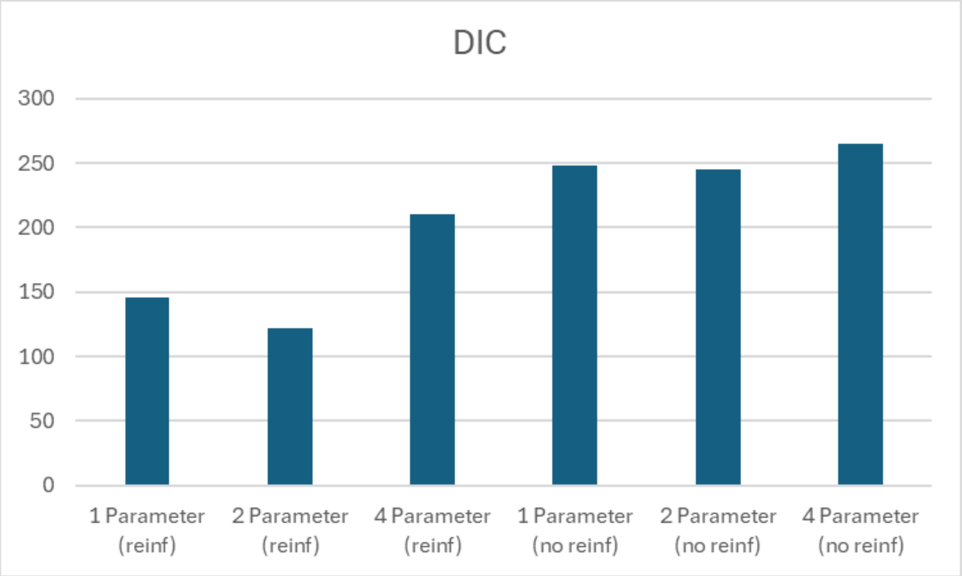

Figure S4: DIC for the different model configurations in the analysis

### Estimation of reproductive numbers

We consider the **SEIAR** model structure for each strain as described above in equations 1 to 65, which compartmentalises the population:

1. **S**: Susceptible individuals,
2. **E**: Exposed individuals (infected but not yet infectious),
3. **I**: Infectious individuals (cases),
4. **A**: Asymptomatic individuals (infectious through ongoing viral shedding but showing no symptoms),
5. **R**: Recovered individuals.

As explained above, the model incorporates seasonality through a time-dependent transmission rate $\beta(t)$, which captures periodic fluctuations in transmission (e.g., due to environmental or behavioural factors). Thus,

- $\beta(t)$ is the seasonally varying transmission rate,
- $\psi$ is the relative infectiousness of asymptomatic individuals compared to symptomatic individuals.

And we use the next generation matrix approach as described by Diekmann et al. through following steps:

1. Identify Infected Compartments: The infected compartments are *E*, *I*, and *A*.
2. Construct the Jacobian Matrices:
   - Define matrix *F* of new infections:

$$F = \begin{matrix} 0 & \beta(t)S & \psi\beta(t)(S+R) \\ 0 & 0 & 0 \\ 0 & 0 & 0 \end{matrix} \tag{70}$$

   - Define matrix *V* for transitions between compartments:

$$V = \begin{matrix} \varepsilon & 0 & 0 \\ -\varepsilon & \theta & 0 \\ 0 & 0 & \eta \end{matrix} \tag{71}$$

where:

- $\varepsilon$ is the rate at which exposed individuals become infectious,
- $\theta$ is the recovery rate for symptomatic individuals,
- $\eta$ is the recovery rate for asymptomatic individuals.

3. Compute the Next Generation Matrix (NGM):

$$\mathrm{K} = \mathrm{FV} - 1 \tag{72}$$

4. Calculate *R*0: We find R0 for each strain as the spectral radius (dominant eigenvalue) of the NGM:

$$R0 = \rho(K) \tag{73}$$

### Further analysis

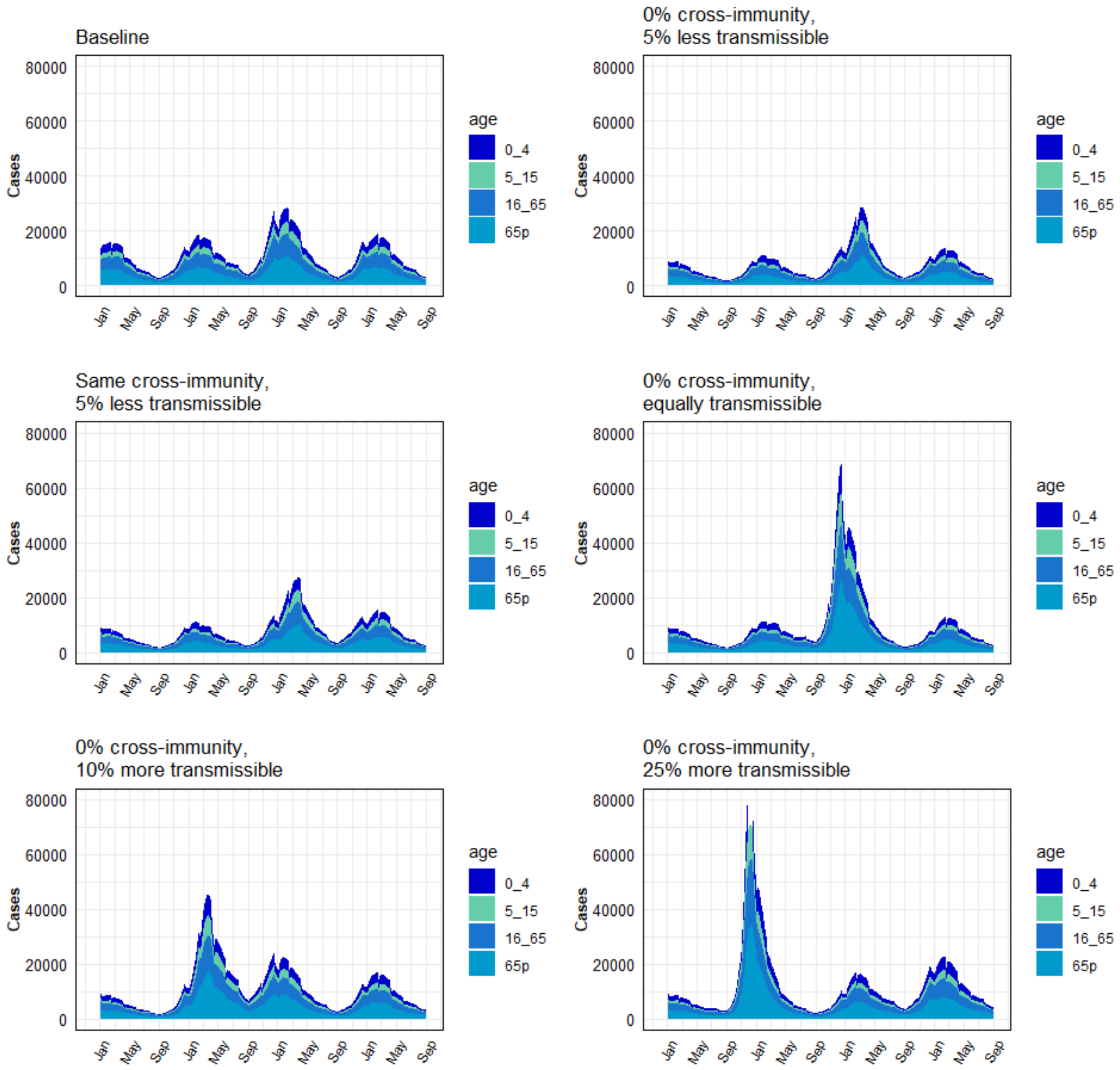

FigS5: Simulated emergence of a novel GII.4 variant: All panels show the average number of modelled Norovirus episodes, stacked by **age**, over the simulation time. An emerging GII.4 variant is introduced in January at the beginning of the simulation. Different levels of GII cross-immunity and transmissibility are explored for the new variant.
